# Research and Diagnostic Algorithmic Rules (RADAR) and RADAR plots for the first episode of major depressive disorder: effects of childhood and recent adverse experiences on suicidal behaviors, neurocognition and phenome features

**DOI:** 10.1101/2022.12.18.22283606

**Authors:** Michael Maes, Abbas F Almulla

**Affiliations:** Department of Psychiatry, Faculty of Medicine, Chulalongkorn University, Bangkok, Thailand; Cognitive Fitness and Technology Research Unit, Faculty of Medicine, Chulalongkorn University, Bangkok, Thailand; Department of Psychiatry, Medical University of Plovdiv, Plovdiv, Bulgaria; Research Institute, Medical University Plovdiv, Plovdiv, Bulgaria; Medical Laboratory Technology Department, College of Medical Technology, The Islamic University, Najaf, Iraq

**Keywords:** psychiatry, mood disorders, maJor depression, neurmmmune, oxidative stress, precision medicine models

## Abstract

This study was performed to ascertain whether a) a valid precision model and valid Research and Diagnostic Algorithmic Rules (RADAR) scores can be computed in patients experiencing first-episode depression; and b) adverse childhood experiences (ACEs) and negative life events (NLEs) are associated with suicidal behavior (SBs), cognitive impairment, and phenome RADAR scores.

This study recruited 90 patients with major depressive disorder (MDD) in an acute phase, of whom 71 showed a first-episode MDD (FEM), and 40 controls. We constructed RADAR scores for ACEs, NLEs encountered the last year, SBs and severity of depression, anxiety, chronic fatigue and physiosomatic symptoms using the Hamilton Depression and Anxiety Rating scales and the Fibro-Fatigue scale.

Partial least squares analysis showed that in FEM, one latent vector (labelled the phenome of FEM) could be extracted from depressive, anxiety, fatigue, physiosomatic, melancholia, and insomnia symptoms, SBs, and cognitive impairments. The latter were conceptualized as a latent vector extracted from the Verbal Fluency Test, Mini Mental State Examination, and ratings of memory and judgment, indicating a generalized cognitive decline (G-CoDe). We found that 60.8% of the variance in the FEM phenome was explained by the cumulative effects of NLEs and ACEs, in particular emotional neglect and, to a lesser extent, physical abuse, and by the interaction between ACEs and physical abuse, whereby the latter attenuates the effects of NLEs.

In conclusion, not the binary diagnosis of MDD (DSM/ICD) should be used in research and clinical settings but the RADAR scores and plots constructed here.

## Introduction

Recent research has shown that there are no valid models of major depressive disorder (MDD) and that there is no replicable and cross-validated model that can be used as an outcome variable in biomarker research at this time (Maes, 2022; Maes and Stoyanov, 2022). When discussing depression, it seems as though psychiatrists cannot understand one another and speak different languages. Different concepts of models (from folk psychology to molecular psychiatry) and subtypes or subclasses (MDD, melancholia, recurrent depressive disorder, dysthymia, double depression, reactive depression, vital depression, treatment resistant depression) rule in chaos (Maes, 2022; 2023). As a result, research on MDD is plagued by severe noise, resulting in a cacophony of models, labels and subtypes without a solid consensus among psychiatrists.

In addition, folk psychologists and sociologists ascertain that depression is a “boundary experience” and that “psychiatry transformed normal sorrow into depressive disorder and contributed to the medicalization of feeling blue, grieving, demoralization, and sadness” (van Os and Kohne, 201; Kohne and Van Os, 2021; Summerfield, 2006; Frances, 2013). Consequently, in psychiatric research, severe medical phenotypes and common emotional distress responses are grouped together, resulting in an entirely heterogeneous MDD study sample (Maes, 2022; 2023). The Western gold standard to diagnose MDD using DSM (American Psychiatric Association, 2013) or ICD (World Health Organization, 2004) criteria exacerbates this chaos (Maes, 2022; 2023). Indeed, the DSM and ICD definitions of mood disorders lack psychiatrist consensus, are unreliable and invalid, leading to misdiagnoses and misclassifications (Maes, 2022; Maes and Stoyanov, 2022). In addition, the top-down dogmatism of the DSM/ICD case definitions preclude inductive (because top-down) and deductive (because it is indisputable unless by the same group of professionals) reworking of the criteria (Maes, 2022). As a result, their use as an explanatory or mediating variable in statistical analyses is not only conceptually flawed, but also results in a multitude of errors and inaccurate conclusions (Maes, 2022; 2023).

Recently, we have created a new supervised and unsupervised machine learning clinimetrics approach called “precision nomothetic psychiatry”, which allows us to build new pathway phenotypes and endophenotype classes (Maes et al., 2021; 2022a; Maes, 2022; 2023 Simeonova et al., 2021; Stoyanov and Maes, 2021). With the help of those methods, we were able to develop i) bottom-up, data-driven nomothetic psychiatry models of MDD, ii) new pathway phenotypes of MDD in the form of phenome (the symptomatome of MDD) scores and a recurrence of illness (ROI) index, based on the recurrence of lifetime suicide ideation (SI) and attempts (SA) and depressive episodes, and iii) a new endophenotype class in the form of Major Dysmood Disorder (MDMD) as opposed to simple DMD (SDMD) (Maes et al., 2021; 2022a; Maes, 2022; 2023; Simeonova et al., 2021; Stoyanov and Maes, 2021). The latter is a qualitatively distinct form of MDD characterized by more adverse childhood experiences (ACEs), a higher prevalence of lifetime SI and SA and recent suicidal behaviors (SBs), a higher ROI, more cognitive impairments, a more severe phenome, and the presence of multiple adverse outcome pathways (AOPs), namely activated nitro-oxidative stress, inflammatory and autoimmune pathways (Maes et al., 2021; 2022a; Maes, 2022; 2023; Simeonova et al., 2021).

Previous studies by our team in MDD have established that ACEs are causal factors in ROI, the phenome and lifetime and current SBs (Maes et al., 2018; 2022b; Maes, 2022; 2023). While genetics and adverse outcome pathways play a key role in the development of MDD, ACEs, and negative life events (NLEs) in the year before to the onset of depression also contribute, especially in the early stages of illness (Paykel, 2003; Kendler et al., 1999; Maes et al. 2018; 2022b).

Recently, we also provided algorithms for computing Research and Diagnostic Algorithm Rule (RADAR) scores for ACEs, ROI, lifetime and current SBs, phenome scores, and the lifetime trajectory (which is a composite of ACEs, ROI, SB, and phenome scores) of mood disorder patients (Maes, 2022; 2023). We showed also how to plot all of those different features of depression as RADAR scores in a two-dimensional RADAR or spider graph, whereby the patient’s data can be visualized, much like a fingerprint, which aids in quickly evaluating the patient’s features. By consolidating the multiple RADAR scores into one simple graph, our method demonstrates how simplistic and minimal the DSM-5 and ICD diagnoses really are by reducing all features into an unreliable, binary MDD diagnosis. Specifically, we argued that clinicians and psychiatric research should always use the derived RADAR scores reflecting ACEs, ROI, SB, neurocognitive, phenome and lifetime trajectory scores, as well as the new endophenotype class MDMD, rather than relying on invalid binary diagnoses (Maes, 2022; 2023). ROI is the most important factor in this precision model because it determines the severity of current SBs and the phenome in both the acute and partially remitted phases of depression (Maes et al., 2019; 2021; 2022a). Our models were developed on patients in the acute and remitted phases of recurrent depression in Brazil and Thailand and who had a wide range of ROI scores.Open questions include whether NLEs combined with ACEs increase the risk of developing new-onset MDD and whether our nomothetic model and RADAR scores can be used in first-episode MDD in other countries and cultures.

Hence, this study was carried out to ascertain whether (a) a valid nomothetic model and valid RADAR scores (excluding ROI scores) can be computed in Iraqi patients experiencing their first depressive episode; b) combined effects of ACE and NLEs increase vulnerability to new-onset depression; and c) whether there are any differences in RADAR scores between first- and second-episode MDD.

## Methods

### Participants

In the present case-control study, 90 patients with a major depressive episode (MDD) were recruited between February 2021 and March 2022 at the psychiatry unit of Al-Hakeem Hospital in Al-Najaf region, Iraq. A senior psychiatrist diagnosed MDD based on DSM-5 (APA, 2013) criteria and selected 71 patients with MDD, more specifically first-episode MDD, moderate or severe without psychotic symptoms, and 19 individuals with recurrent MDD, moderate or severe without psychotic features. The primary focus of the current study was to build a nomothetic model and construct RADAR scores and graphs in first-episode patients. A secondary aim was to compare the RADAR scores among first-episode and second-episode patients. All patients with MDD were in the acute phase of the disease, and none exhibited complete or partial remission. The mean (SD) duration of illness for patients with first episode MDD was 2.5 (±0.3) months. Twenty-seven of the 71 first-episode patients were drug naive, whereas all second-episode patients received medication for at least 3 weeks: 42 patients were given fluoxetine, 10 were given amitriptyline, 8 were given escitalopram, 12 were given mirtazapine, and 9 were given olanzapine. The same senior psychiatrist also recruited forty apparently healthy controls from the same catchment area as medical staff or their friends and patients’ friends. Patients and controls were excluded for any other axis-1 DSM-5 disorders, such as autism and ASD, dysthymia, schizophrenia, bipolar disorder, substance use disorders (all except nicotine dependence), major anxiety disorders, including generalized anxiety disorder and panic disorder, post-traumatic stress disorder, and obsessive-compulsive disorder. In addition, controls with a lifetime diagnosis of MDD or a family history of depression, bipolar disorder, substance use disorders, or psychosis were excluded. We also excluded pregnant and lactating women and subjects with a) neurodegenerative or neuroinflammatory disorders, including stroke, multiple sclerosis, and Parkinson’s and Alzheimer’s disease; b) chronic liver and kidney disorders; and c) (auto)immune disease, including psoriasis, rheumatoid arthritis, inflammatory bowel disease, cancer, type 1 diabetes, scleroderma, moderate and critical COVID-19, and rheumatoid arthritis. In addition, subjects treated with immunosuppressive or immunomodulatory drugs or therapeutic doses of antioxidants or omega-3 supplements were ineligible for participation.

All participants provided written informed consent prior to the enrollment in the study. The study complied with international and Iraqi laws governing ethics and privacy. According to the International Guidelines for the Protection of Human Subjects of the Declaration of Helsinki, the study was approved by the ethics committee (IRB) of the College of Medical Technology, The Islamic University of Najaf, Iraq (Document No. 18/2021).

### Measurements

The senior psychiatrist conducting the study collected demographic (marital, occupational, educational) and clinical data (duration of the index episode, age at onset, prior COVID infection and severity) using a semi-structured interview. He utilized DSM-5 diagnostic criteria (APA, 2013) to identify MDD patients and exclude those with other axis-1 diseases. We assessed ACEs using the Adverse Childhood Experience (ACE) Questionnaire (Felitti et al., 1998), which assesses 10 major abuse, neglect and household dysfunction domains as present or not present, including ACEQ1: emotional abuse; ACEQ2: physical abuse; ACEQ3: sexual abuse; ACEQ4: emotional neglect; ACEQ5: physical neglect; ACEQ6: divorce; ACEQ7: violent behavior; ACEQ8: substance abuse; ACEQ9: mental illness; and ACEQ10: incarcerated relative. Negative Life Events (NLE) the year prior to the onset of depression were assessed using the Negative Life Events scale (Cohen, Tyrrell et al., 1993) and we considered the following items to be relevant namely serious accidents, death of a family member or close relative, divorce or separation, seeing fights, abuse or violent crime, trouble with the police, and a member of the family sent to jail. Suicidal behaviors were assessed using two items of the Columbia Suicide Severity Rating Scale (C-SSRS), namely a) weekly frequency of suicidal ideation the last three months; and b) number of suicidal attempts the year previous to inclusion in the study (Posner et al., 2011). Cognitive functioning was assessed using the Verbal Fluency Test (VFT) (Shao et al., 2014) to assess word fluency and semantic memory; Mimi Mental State Examination (MMSE) (Folstein et al., 1975) and the Clinical Dementia Rating (CRC) (Morris, 1993) which assesses 6 domains on a 0-3 point scale, namely memory, orientation, judgement and problem solving, community affairs, home & hobbies, and personal care. Nevertheless, we did not use the CDR as proposed by Morris (1993), but used to sum on memory, orientation and judgement coupled with the VFT and MMSE to examine whether we could extract one meaningful and validated principal component (PC) (see below). We also intended to examine the other 3 items (community, grooming and hobbies) as possibly contributing to the phenome scores of MDD. However, we could not find any relevance of these measurements. The severity of depression and anxiety was assessed using the Hamilton Depression (HAMD) and Anxiety (HAMA) Rating Scales (Hamilton, 1959; 1960). We used the FibroFatigue scale (Zachrisson et al. 2002) to assess severity of fibromyalgia- and chronic fatigue-like and physiosomatic symptoms.

### Statistics

We utilized analysis of variance (ANOVA) or the Mann-Whitney U test to compare continuous variables and analysis of contingency tables (χ^2^-test) to compare nominal variables between groups. Using Pearson’s product moment correlation coefficients, we examined the links between scale variables. Using multiple regression analysis, the effects of explanatory variables (ACEs, NLEs, age, sex, and education) on dependent variables (e.g. symptomatome and phenome scores) were examined. In addition, we used a forward stepwise automatic regression method using p-values of 0.05 to-enter and 0.06 to remove for inclusion and exclusion in the final regression model. We generated the standardized β coefficients with t-statistics and exact p-values for each of the explanatory variables in the final regression models, in addition to F statistics (and p values) and total variance (R^2^ or partial eta squared used as effect size) explained by the model. Collinearity and multicollinearity were investigated using tolerance (cut-off value 0.25), the variance inflation factor (cut-off value >4), and the condition index and variance proportions from the collinearity diagnostics table. The White and modified Breusch-Pagan tests were used to verify the presence of heteroskedasticity. All above tests were two-tailed, and an alpha value of 0.05 was deemed statistically significant. In order to normalize the distribution of the data some variables were entered as transformation, including logarithmic or rank-based inversed normal (RINT) transformations.

We used principal component (PC) analysis to check whether a set of MDD features could be reduced to one meaningful PC. To be acknowledged as a validated PC, the first PC must account for >50% of the variance in the data, all loadings on this factor must be > 0.7, and the factoriability of the correlation matrix must be satisfactory, as determined by the Kaiser-Meyer-Olkin (KMO) test (KMO should be > 0.6), the Bartlett’s test of sphericity (p should be < 0.05) and the anti-image matrix which should be sufficient. We used two-step cluster analysis to identify relevant groups of individuals based on the features of the disease. When the silhouette measure of cohesiveness and separation was more than 0.5, the cluster-generated solution was deemed appropriate. All preceding machine learning analyses were conducted using Windows version 28 of the IMB-SPSS application.

Path analysis using partial least squares (PLS) (Ringle et al., 2014) was used to predict the final outcome (output) variables, namely the phenome of mood disorders, using a set of independent (input) variables including ACEs, NLEs and neurocognitive impairments. Additionally, the model accounts for mediated effects (e.g. the effects of ACEs on the phenome are mediated by the G-CoDe). Variables were either entered as single indicators or as latent vectors (e.g. a factor extracted from all symptom domains). Complete PLS path analysis was only performed if the inner and outer models met the following predetermined quality criteria: a) the latent vectors of the outer models demonstrate high convergent and construct validity as indicated by Cronbach’s alpha > 0.7, composite reliability > 0.8, rho A > 0.8, and high loadings (>0.7) at p<0.0001 of the indicators of the latent vectors; and b) the overall model fit, namely the standardized root mean square (SRMR) is < 0.08. PLSPredict and the cross-validated predictive ability test (CVPAT) were used to evaluate the replicability of the final PLS model. Q^2^ values were used to estimate whether the model’s prediction error is significantly smaller than the prediction error of the naive and linear regression model benchmarks. In addition, we employed Confirmatory Tetrad Analysis (CTA) to ensure that the latent constructs were not incorrectly specified as a reflective model. Permutation and Multi-Group Analysis (MGA) were utilized to investigate whether predifined groups (including men versus women, smokers versus non-smokers, drug-naive versus medicated) show significant differences in parameter estimates (SmartPLS, 2022; Henseler et al., 2016; Cheah et al., 2020; Sarsyedt et al., 2011) and to delineate whether the models originate from a common population. Invariance Assessment of Composite Models (MICOM) was utilized to evaluate “configural and compositional invariance, and the equality of composite mean values and variances” (SmartPLS, 2022; Henseler et al., 2016; Cheah et al., 2020; Sarsyedt et al., 2011). Using the Heterotrait - Monotrait (HTMT) ratio with a cutoff value of > 0.85, the discriminant validity of the constructs was determined. If all the previously mentioned model fit quality data met the predetermined criteria, we conducted a complete PLS path analysis with 5,000 bootstrap samples, calculated path coefficients (with exact p-values), specific indirect, total indirect (i.e., mediated) and total effects.

## Results

### Construction of different RADAR scores

Based on our previous publications (Maes, 2022; 2023), we computed several symptom subdomain and RADAR scores as z unit-based composite scores summing up the z scores of different items, namely:

1. The ACE score was computed as the sum of the 10 ACEs because it was impossible to extract validated and replicable PCs from the data. We also entered all single ACE indicators in the analyses (except Q3, Q5 and Q8 which showed virtually no variance).
2. The NLE score was computed as the presence of any NLE item encountered the last year. We have examined how ACEs and NLEs could best be presented, namely separately, an interaction term ACE x NLE, the sum of the different adverse events (AEs) and detected that in ANOVAs the sum of ACEs + NLE (yes/no) was most appropriate (labeled AEs), whereas in regression analysis it was most appropriate to enter the separate ACEs and NLEs scores. Furthermore, the interaction pattern was also significant in regression analyses. Consequently, the adverse experience (AEs) score was computed as the RINT of total ACE + NLE (yes/no). Using a visual binning method, the study sample was divided into three samples, namely a group with few AEs (< −0.59), one with some AEs (< 0.60 – 0.58), and another group with many AEs (> 0.59).
3. The pure depressive domain score was computed as a z based composite score of depressed mood + feelings of guilt + loss of interest (HAMD) + sadness (FF) + depressed mood (HAMA).
4. The pure anxiety domain score was computed as a z-based composite score as the sum of anxious mood + tension + fears + anxiety behavior at interview + anxiety psychological (HAMD). Both the pure depression and depression scores were processed as RINT scores.
5. The pure physiosomatic symptom domain score was computed as a z unit-based composite score based on the sum (z scores) of anxiety somatic + gastrointestinal + genitourinary + hypochondriasis somatic sensory + cardiovascular + gastrointestinal (GIS) + genitourinary + autonomic symptoms + respiratory symptoms (HAMA symptoms) + muscle pain + muscle tension + fatigue + autonomic + gastro-intestinal + headache + malaise (FF scale) + anxiety somatic + somatic gastro-intestinal + general somatic + genital symptoms + hypochondriasis (HAMD).
6. The melancholia domain score was computed as the sum of insomnia late + psychomotor retardation + psychomotor agitation + loss of weight + diurnal variation.
7. The insomnia domain score was calculated as a z unit-based composite score computed as insomnia early + insomnia middle + insomnia late (HAMD) + sleep disorders (FF) + insomnia (HAMA).
8. The subjective cognitive impairment (SCI) score was computed as a z unit-based composite calculated as concentration disorders + memory disturbance (FF scale) + intellectual problems (HAMA) + cognition (HAMD).
9. The phenome1 score was computed as a PC extracted from the above 6 symptom domains. The first PC showed an adequate model fit with AVE=79.21%, Cronbach alpha=0.921, factor loadings that were all > 0.786 (KMO=0.885, Bartlett’s χ2=708.05, df=15, p<0.001).
10. The suicidal behavior (SB) score was computed as a composite score calculated as frequency of suicidal ideation + frequency suicidal attempts + suicidal ideation (HAMD item).
11. The phenome2 score was computed as the first PC score extracted from the 6 symptom domains and the SB score. The first PC showed an adequate model fit with AVE=77.06%, Cronbach alpha=0.930, factor loadings that were all > 0.770 (KMO=0.904, Bartlett’s χ2=826.60, df=21, p<0.001).
12. In accordance with recent findings in schizophrenia that one factor reflecting a generalized cognitive decline (G-CoDe) could be extracted from several cognitive tests results (Maes and Kanchanatawan, 2021), we examined whether one PC could be extracted from different neurocognitive tests in MDD. Indeed, we were able to extract a G-Code construct from MMSE, VFT and the sum of 3 CDR item scores (memory + orientation + judgement). This first PC showed an AVE=63.65%, Cronbach alpha=0.691, and factor scores > 0.763 (KMO=0.666, Bartlett’s χ2=62.98, df=3, p<0.001).
13. The phenome3 score was computed as a PC score extracted from all 6 symptom domains, SBs and G-CoDe. This first PC showed an adequate model fit with AVE=75.84%, Cronbach alpha=0.939, factor loadings that were all > 0.776 (KMO=0.918, Bartlett’s χ2=945.31, df=28, p<0.001).
14. The ROI score in the total study group (thus with second-episode patients included) was computed as the RINT transformation of a composite score built using frequency of suicidal ideation, frequency of suicidal attempts and number of episodes (the ROI score can only be used when analyzing the total study group).
15. The lifetime trajectory score was assessed as the first PC score extracted from AEs, SBs, G-CoDe and phenome1 scores. This first PC showed an adequate model fit with AVE=70.59%, Cronbach alpha=0.860, and factor loadings > 0.794 (KMO=0.821, Bartlett’s χ2=198.21, df=6, p<0.001).

### Features of study groups with low, some and many AEs

**Table 1** shows the socio-demographic and clinical variables in subjects divided into those with few, some and many AEs. There were no significant differences in age, sex ratio, BMI, education, married status, TUD and prior COVID-19 infection between the three study groups. There was a significant association between this division and the diagnosis of first-episode major depression. Subjects with some and many AEs showed lowered G-CoDe and SCIs scores than subjects with few AEs. There were significant differences in SBs, all symptom domains other than SCIs, and phenome2 and phenome3 scores between the three study groups with increasing scores from few → some → many AEs. Subjects with NLEs showed a significantly higher mean (SD) number of ACEs (1.73 ±0.98) as compared with those without NLEs (0.98 ±109) (Mann-Whithney U test: <0.001).

**Table 1.**
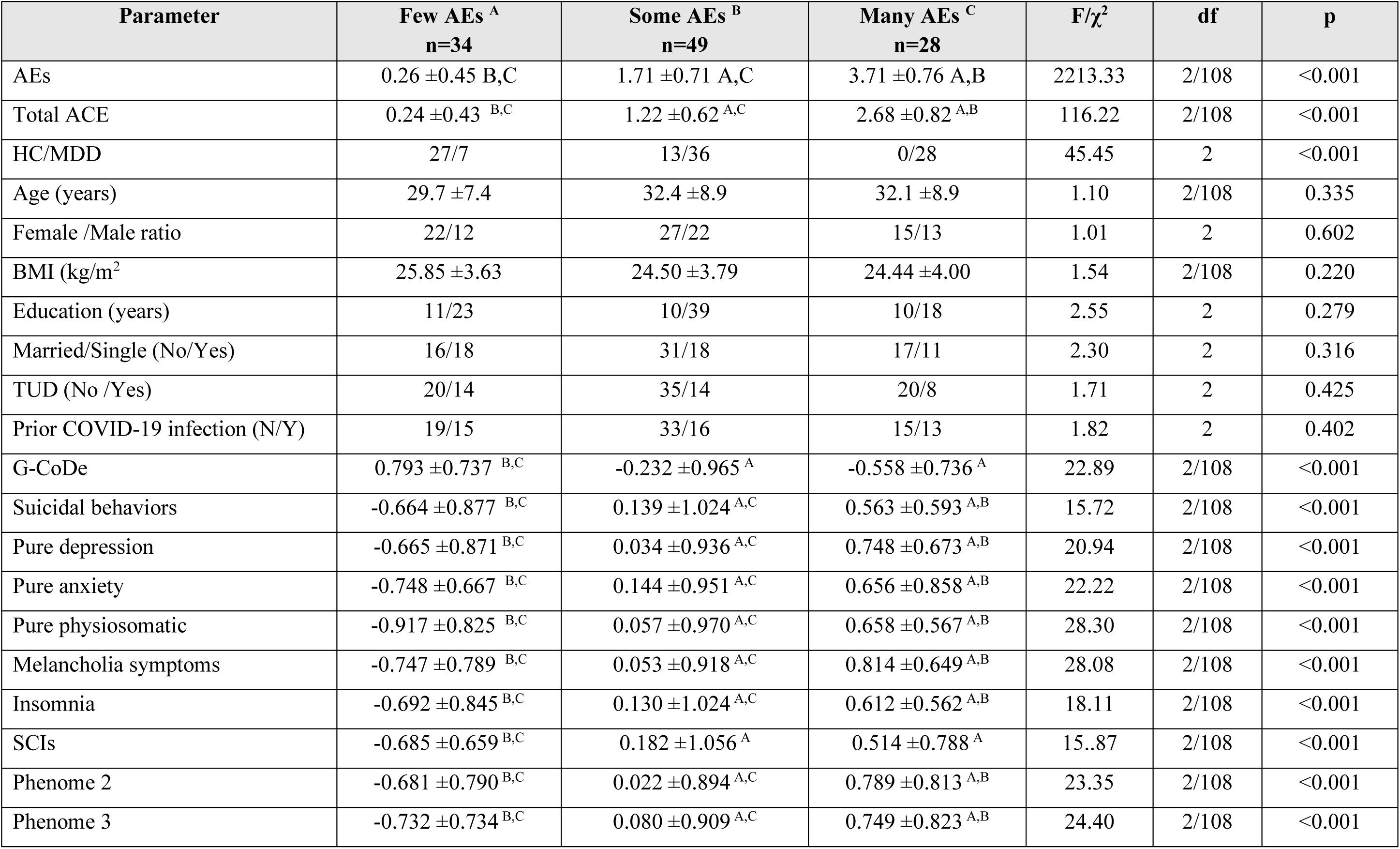
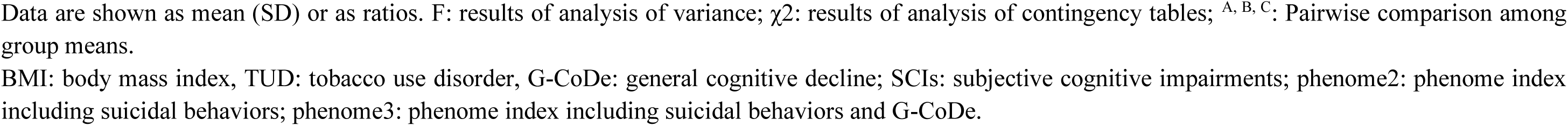
Socio-demographic and clinical variables in subjects divided into those with few, some and more adverse experiences (AEs)

| Parameter | Few AEs <sup>A</sup><br>n=34 | Some AEs <sup>B</sup><br>n=49 | Many AEs <sup>C</sup><br>n=28 | F/$\chi^2$ | df | p |
| --- | --- | --- | --- | --- | --- | --- |
| AEs | 0.26 ±0.45 B,C | 1.71 ±0.71 A,C | 3.71 ±0.76 A,B | 2213.33 | 2/108 | <0.001 |
| Total ACE | 0.24 ±0.43 <sup>B,C</sup> | 1.22 ±0.62 <sup>A,C</sup> | 2.68 ±0.82 <sup>A,B</sup> | 116.22 | 2/108 | <0.001 |
| HC/MDD | 27/7 | 13/36 | 0/28 | 45.45 | 2 | <0.001 |
| Age (years) | 29.7 ±7.4 | 32.4 ±8.9 | 32.1 ±8.9 | 1.10 | 2/108 | 0.335 |
| Female /Male ratio | 22/12 | 27/22 | 15/13 | 1.01 | 2 | 0.602 |
| BMI (kg/m<sup>2</sup>) | 25.85 ±3.63 | 24.50 ±3.79 | 24.44 ±4.00 | 1.54 | 2/108 | 0.220 |
| Education (years) | 11/23 | 10/39 | 10/18 | 2.55 | 2 | 0.279 |
| Married/Single (No/Yes) | 16/18 | 31/18 | 17/11 | 2.30 | 2 | 0.316 |
| TUD (No /Yes) | 20/14 | 35/14 | 20/8 | 1.71 | 2 | 0.425 |
| Prior COVID-19 infection (N/Y) | 19/15 | 33/16 | 15/13 | 1.82 | 2 | 0.402 |
| G-CoDe | 0.793 ±0.737 <sup>B,C</sup> | -0.232 ±0.965 <sup>A</sup> | -0.558 ±0.736 <sup>A</sup> | 22.89 | 2/108 | <0.001 |
| Suicidal behaviors | -0.664 ±0.877 <sup>B,C</sup> | 0.139 ±1.024 <sup>A,C</sup> | 0.563 ±0.593 <sup>A,B</sup> | 15.72 | 2/108 | <0.001 |
| Pure depression | -0.665 ±0.871 <sup>B,C</sup> | 0.034 ±0.936 <sup>A,C</sup> | 0.748 ±0.673 <sup>A,B</sup> | 20.94 | 2/108 | <0.001 |
| Pure anxiety | -0.748 ±0.667 <sup>B,C</sup> | 0.144 ±0.951 <sup>A,C</sup> | 0.656 ±0.858 <sup>A,B</sup> | 22.22 | 2/108 | <0.001 |
| Pure physiosomatic | -0.917 ±0.825 <sup>B,C</sup> | 0.057 ±0.970 <sup>A,C</sup> | 0.658 ±0.567 <sup>A,B</sup> | 28.30 | 2/108 | <0.001 |
| Melancholia symptoms | -0.747 ±0.789 <sup>B,C</sup> | 0.053 ±0.918 <sup>A,C</sup> | 0.814 ±0.649 <sup>A,B</sup> | 28.08 | 2/108 | <0.001 |
| Insomnia | -0.692 ±0.845 <sup>B,C</sup> | 0.130 ±1.024 <sup>A,C</sup> | 0.612 ±0.562 <sup>A,B</sup> | 18.11 | 2/108 | <0.001 |
| SCIs | -0.685 ±0.659 <sup>B,C</sup> | 0.182 ±1.056 <sup>A</sup> | 0.514 ±0.788 <sup>A</sup> | 15.87 | 2/108 | <0.001 |
| Phenome 2 | -0.681 ±0.790 <sup>B,C</sup> | 0.022 ±0.894 <sup>A,C</sup> | 0.789 ±0.813 <sup>A,B</sup> | 23.35 | 2/108 | <0.001 |
| Phenome 3 | -0.732 ±0.734 <sup>B,C</sup> | 0.080 ±0.909 <sup>A,C</sup> | 0.749 ±0.823 <sup>A,B</sup> | 24.40 | 2/108 | <0.001 |

Data are shown as mean (SD) or as ratios. F: results of analysis of variance; $\chi^2$ : results of analysis of contingency tables; <sup>A, B, C</sup>: Pairwise comparison among group means.
BMI: body mass index, TUD: tobacco use disorder, G-CoDe: general cognitive decline; SCIs: subjective cognitive impairments; phenome2: phenome index including suicidal behaviors; phenome3: phenome index including suicidal behaviors and G-CoDe.

### Correlations between ACEs, AEs and symptom domains

**Table 2** shows the correlation matrix between ACEs and AEs and the symptom domains assessed in controls and first-episode MDD patients. ACEs and AEs were significantly and negatively correlated with G-CoDe and positively with all other symptom domains.

**Table 2.** Correlation matrix between number of adverse childhood experiences (ACEs) alone and combined with negative life events (AEs) and the symptom domains assessed in this study.

| **Variables** | **Total ACE\*** | **AEs\*** | **Total ACE\*\*** | **AEs\*\*** |
| --- | --- | --- | --- | --- |
| G-CoDe | -0.495 | -0.600 | -0.489 | -0.586 |
| Suicidal behaviors | 0.389 | 0.484 | 0.391 | 0.484 |
| Pure depression | 0.515 | 0.559 | 0.502 | 0.548 |
| Pure anxiety | 0.455 | 0.565 | 0.434 | 0.528 |
| Pure physiosomatic | 0.483 | 0.618 | 0.506 | 0.616 |
| Melancholia | 0.510 | 0.597 | 0.534 | 0.617 |
| Insomnia | 0.486 | 0.563 | 0.486 | 0.557 |
| SCIs | 0.412 | 0.473 | 0.365 | 0.423 |
| Phenome 2 | 0.468 | 0.553 | 0.494 | 0.565 |
| Phenome 3 | 0.479 | 0.570 | 0.497 | 0.576 |

All significant at $p < 0.001$ , \*Performed in controls and first episode depressed patients ( $n=111$ ); \*\*Performed in controls and all depressed patients combined ( $n=130$ ); G-CoDe: general cognitive decline; SCIs: subjective cognitive impairments; phenome2: phenome index including suicidal behaviors; phenome3: phenome index including suicidal behaviors and the G-CoDe.

### Multiple regression analysis with phenome features as dependent variables

**Table 3** shows the results of multiple regression analyses with the phenome features as dependent variables and adverse childhood experiences (ACEs), negative life events (NLE) and AEs as explanatory variables while allowing for the effects of socio-demographic data. Model 1 shows that 35.6% of the variance in the G-CoDe was explained by AEs, and that a combination of ACEQ2, ACEQ4, ACEQ6 and NLE explained up to 45.8% of the variance in neurocognitive impairments (model #2). **Figure 1** shows the partial regression of the G-CoDe on AEs. The latter explained (model #3) 27.4% of the variance in SBs, while ACEQ4, ACEQ10 and NLE explained 32.9% of the variance (model #4). **Figure 2** shows the partial regression of the SB score on AEs. **Figure 3** shows the partial regression of the SB score on AEs. We found that 35.2% of the variance in the phenome3 score (model #5) was explained by the regression on AEs and gender (higher in males) and that ACEQQ4, NLE, and ACEQ2 were the best predictors of the phenome2 scores, explaining 37.3% of the variance (model #6). We found that 37.5% of the variance in the phenome3 score (model #7) was explained by the regression on AEs and male sex (all positively associated) and that male sex, ACEQ2, ACEQ4 and NLE predicted the phenome3 scores and explained 42.9% of the variance (model #8). In model # 9, we added the interaction term Q4 X NLE which contributed significantly towards the phenome3 score (inversely associated).

**Figure 1.**
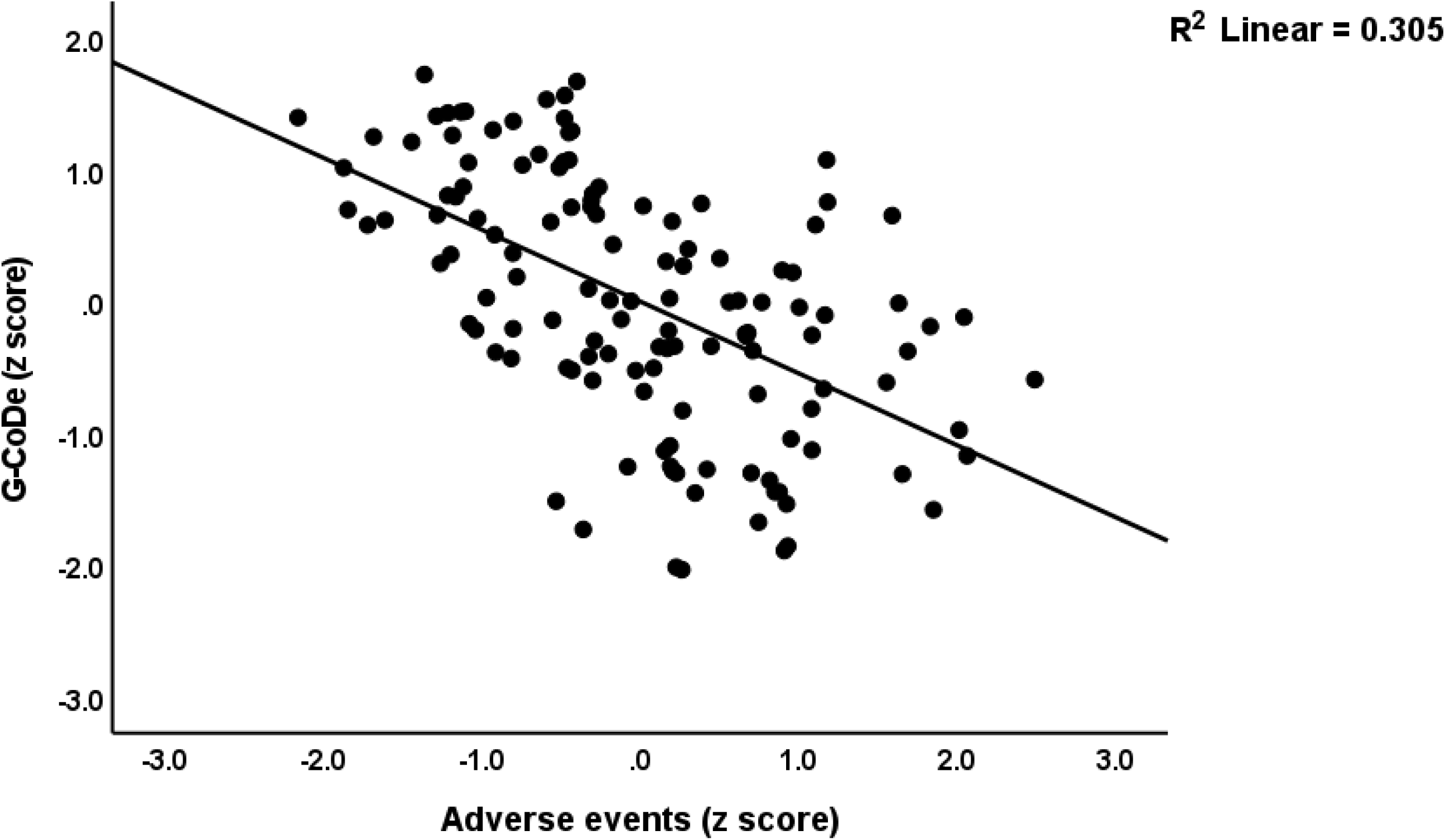
Partial regression of the generalized cognitive decline (G-CoDe) score during the acute phase of first-episode major depression on adverse events.

**Figure 2.**
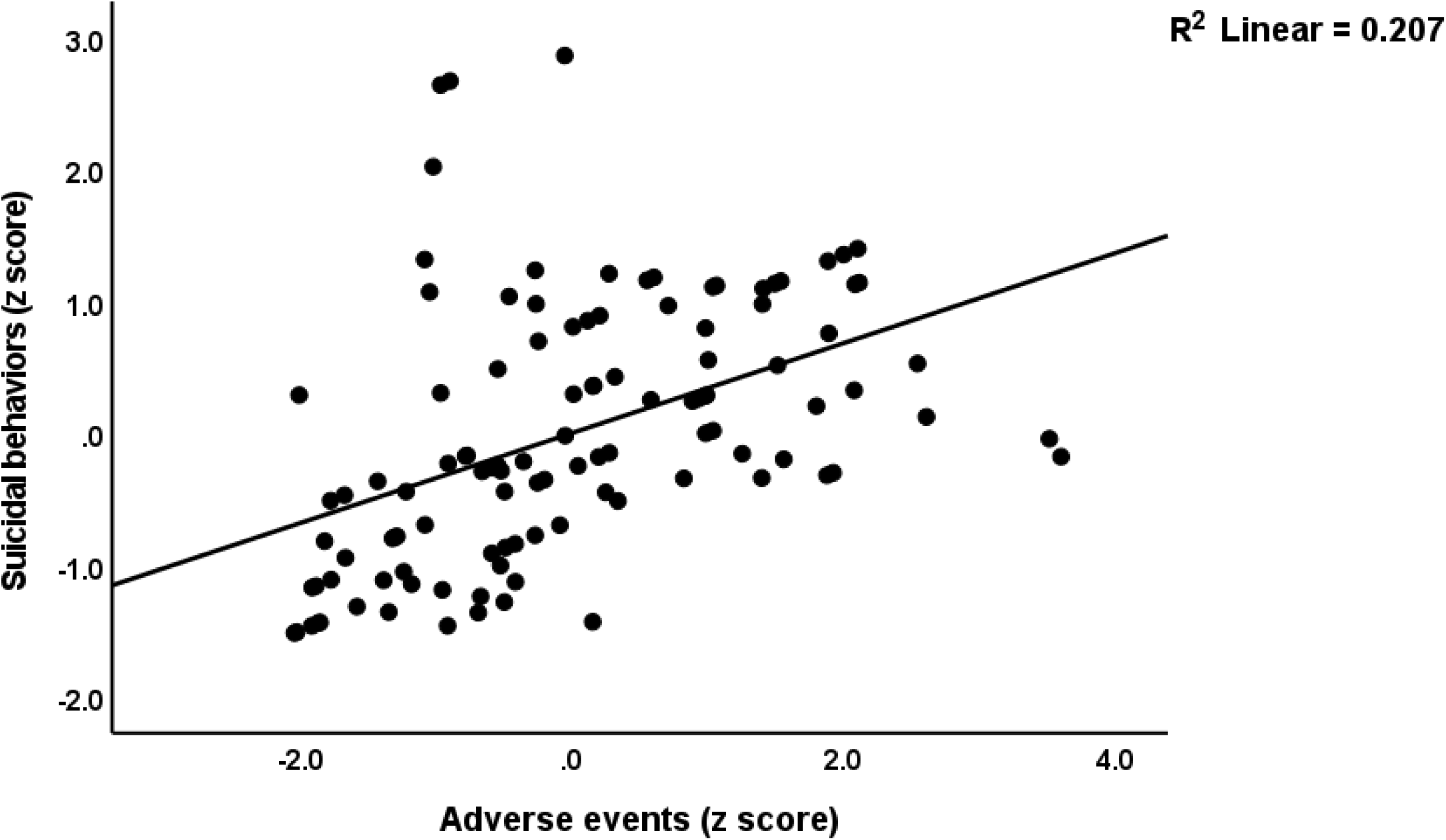
Partial regression of the suicidal behaviors score during the acute phase of first­episode major depression and adverse events.

**Figure 3.**
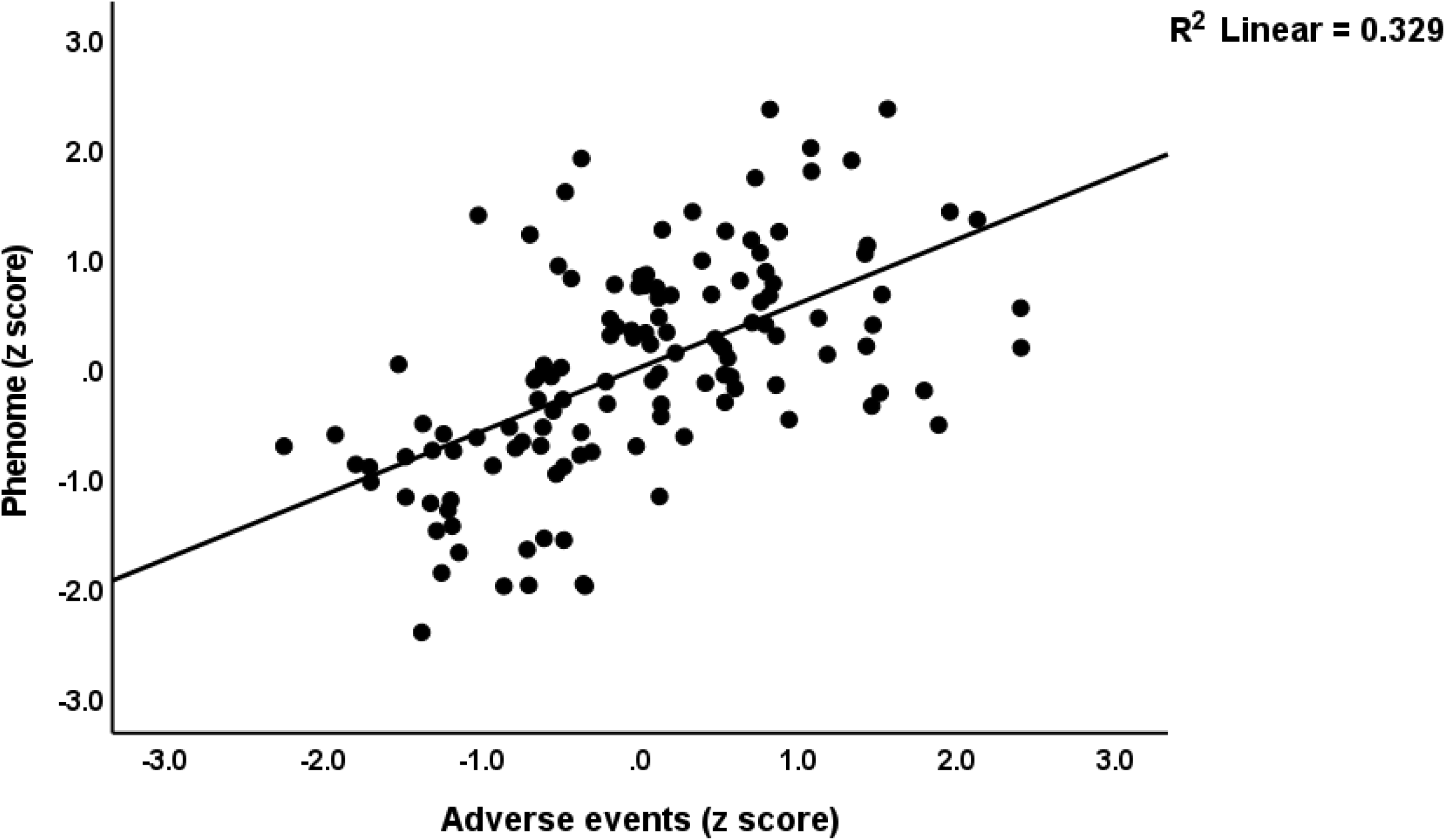
Partial regression of the phenome of the acute phase of first-episode depression on adverse events

**Table 3.** Results of multiple regression analyses with the major symptom domains as dependent variables and adverse childhood experiences (ACEs) and negative life events (NLE) as explanatory variables while allowing for the effects of socio-demographic data.

| Dependent variables | Explanatory variables | B | t | p | F model | df | p | R<sup>2</sup> |
| --- | --- | --- | --- | --- | --- | --- | --- | --- |
| G-CoDe | **Model#1** |  |  |  | 60.19 | 1/109 | <0.001 | 0.356 |
|  | AEs | -0.596 | -7.76 | <0.001 |  |  |  |  |
| G-CoDe | **Model#2** |  |  |  | 22.36 | 4/106 | <0.001 | 0.458 |
|  | ACEQ4 | -0.402 | -5.21 | <0.001 |  |  |  |  |
|  | NLE | -0.307 | -3.94 | <0.001 |  |  |  |  |
|  | ACEQ2 | -0.220 | -3.01 | 0.003 |  |  |  |  |
|  | ACEQ6 | -0.163 | -2.22 | 0.028 |  |  |  |  |
| Suicidal behaviors | **Model#3** |  |  |  | 41.15 | 1/109 | <0.001 | 0.274 |
|  | AEs | 0.524 | 6.42 | <0.001 |  |  |  |  |
| Suicidal behaviors | **Model#4** |  |  |  | 17.50 | 3/107 | <0.001 | 0.329 |
|  | ACEQ4 | 0.328 | 3.79 | <0.001 |  |  |  |  |
|  | NLE | 0.306 | 3.60 | <0.001 |  |  |  |  |
|  | ACEQ10 | 0.182 | 2.26 | 0.026 |  |  |  |  |
| Phenome 2 | **Model#5** |  |  |  | 29.31 | 2/108 | <0.001 | 0.352 |
|  | AEs | 0.574 | 7.41 | <0.001 |  |  |  |  |
|  | Sex | 0.165 | 2.13 | 0.013 |  |  |  |  |
| Phenome 2 | **Model#6** |  |  |  | 21.19 | 3/107 | <0.001 | 0.373 |
|  | ACEQ4 | 0.399 | 4.84 | <0.001 |  |  |  |  |
|  | NLE | 0.278 | 3.38 | 0.001 |  |  |  |  |
|  | ACEQ2 | 0.168 | 2.17 | 0.033 |  |  |  |  |
| Phenome 3 | **Model#7** |  |  |  | 32.43 | 2/108 | <0.001 | 0.375 |
|  | AEs | 0.589 | 7.74 | <0.001 |  |  |  |  |
|  | Sex | 0.185 | 2.43 | 0.017 |  |  |  |  |
| Phenome 3 | **Model#8** |  |  |  | 19.89 | 4/106 | <0.001 | 0.429 |
|  | ACEQ4 | 0.385 | 4.84 | <0.001 |  |  |  |  |
|  | NLE | 0.287 | 3.64 | <0.001 |  |  |  |  |
|  | ACEQ2 | 0.227 | 3.00 | 0.003 |  |  |  |  |
|  | Sex | 0.173 | 2.30 | 0.023 |  |  |  |  |
| **Phenome 3** | **Model#9** |  |  |  | 17.45 | 5/105 | <0.001 | 0.454 |
|  | Sex | 0.174 | 2.35 | 0.021 |  |  |  |  |
|  | NLE | 0.438 | 4.23 | <0.001 |  |  |  |  |
|  | ACEQ2 | 0.178 | 2.29 | 0.024 |  |  |  |  |
|  | ACEQ4 | 0.563 | 5.01 | <0.001 |  |  |  |  |
|  | ACEQ4 X NLE | -0.311 | -2.20 | 0.030 |  |  |  |  |
| **ROI (n=130)** | **Model#10** |  |  |  | 52.39 | 3/126 | <0.001 | 0.555 |
|  | NLE | 0.768 | 9.33 | <0.001 |  |  |  |  |
|  | ACEQ4 | 0.728 | 7.82 | <0.001 |  |  |  |  |
|  | ACEQ4 x NLE | -0.577 | -5.77 | <0.001 |  |  |  |  |

G-CoDe: general cognitive decline; phenome2: phenome index including suicidal behaviors; phenome3: phenome index including suicidal behaviors and G-CoDe; ROI: recurrence of illness based on 2 episodes only; adverse events (AEs); ACE: adverse childhood experiences; NLE: negative life events; Q4: emotional neglect; Q2: physical abuse; Q6: divorce of parents; Q10: imprisonment of a family member.

### Construction of clusters of first-episode patients and RADAR scores

Using two-step cluster analysis with diagnosis of MDD as categorical variable and AEs, phenome 1, 2 and 3 scores and lifetime trajectory as clustering variables. We found three clusters with an adequate silhouette of cohesion and separation of 0.6, namely a first control cluster (n=40), and two patient clusters one with 42 (cluster 2) and another with 29 patients (cluster 3). **Figure 4** shows the features of the three clusters. ANOVA showed that all feature scores were significantly higher in cluster 3 than in cluster 2, except ACEs (p=0.066), AEs (p=0.81), SBs (p=0.176), and sleep disorders (p=0.888). Accordingly, cluster 3 was labeled major dysmood disorder (MDMD) and cluster 2 simple dysmood disorder (SDMD).

**Figure 4.**
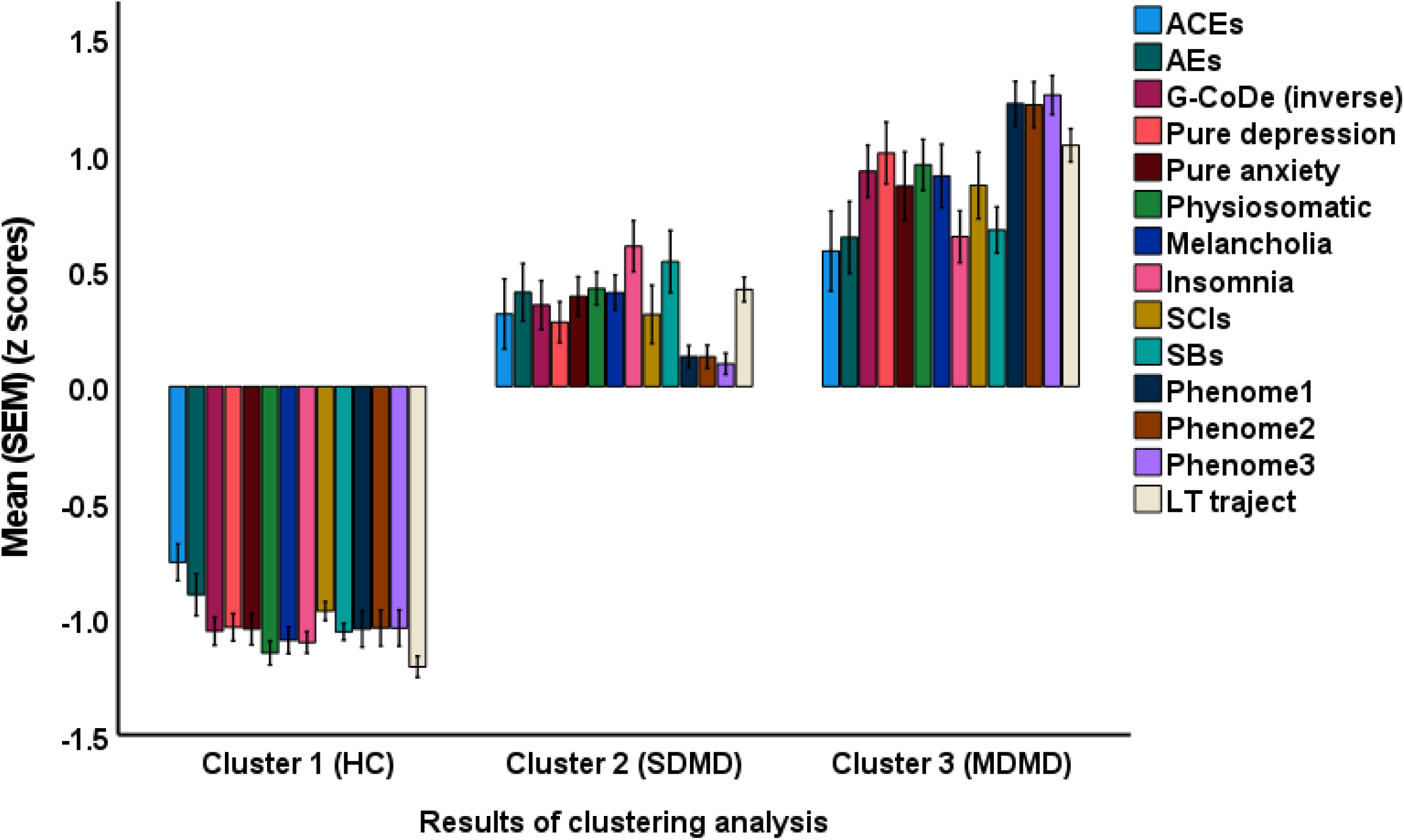
Results of cluster analysis discriminating the acute phase of first-episode major depression into two clusters, one MDMD: major dysmood disorder and SDMD: simple dysmood disorder. ACE: adverse childhood experiences; AEs: adverse events; G-CoDe: generalized cognitive decline; SCIs: subjective cognitive impairments; SBs: suicidal behaviors; LT traject: lifetime trajectory

The RADAR plot in **Figure 5** displays the RADAR scores for one cluster 2 patient with SMDM and one cluster 3 patient with MDMD. The common center point, which was established as the mean value of all feature scores of healthy controls set to 0, is shown in the center of the graph as well as the relative position of the feature scores of the two patients. The RADAR graph provides 14 RADAR or feature scores displayed on 14 radial axes each corresponding to a feature. The latter are ordered along the lifetime trajectory of the individuals starting with ACEs, then AEs, G-CoDe, symptomatome domain features, SBs, phenome scores and finally the lifetime trajectory score. This is important to evaluate the differences in the area shape in the RADAR chart among individuals or groups. This score is expressed in z scores (with mean of healthy controls set as zero) and, thus, shows the difference in scores of both patients in standard deviations versus controls. The radial axes in the RADAR graph are joined in the middle of the figure (zero feature scores of the controls) and are joined by angular axes which divide the graph into grids which show the variation in feature ratings of the two subjects versus normal controls. This graph shows that the RADAR chart of both patients is quite different in particular the ACE, AE, depression, anxiety, physiosomatic, SB, all phenome and lifetime trajectory scores. **Figure 6** shows the RADAR graphs for two other MDD patients categorized as MDMD.

**Figure 5.**
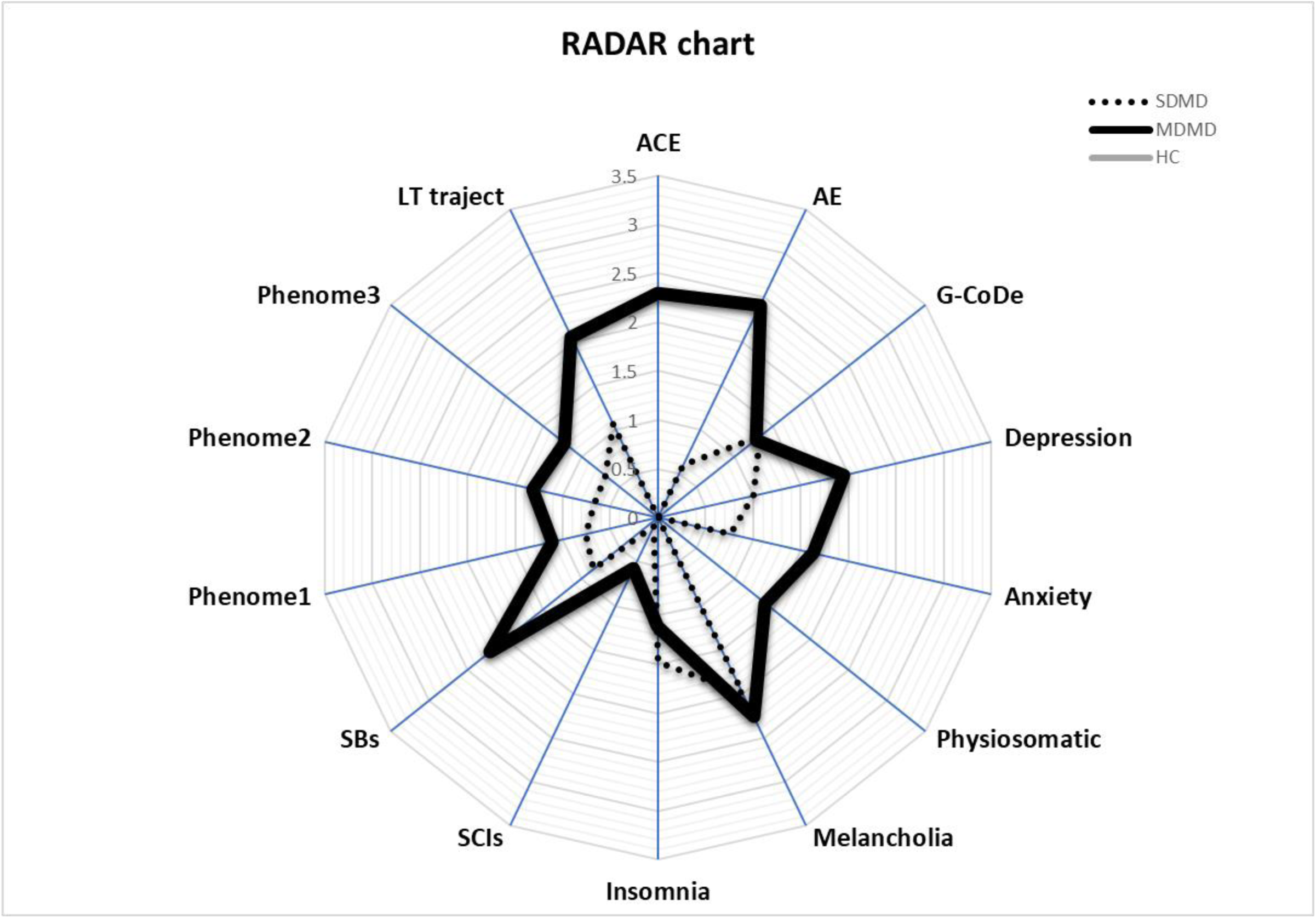
Radar or spider plot displaying the Research and Diagnostic Algorithmic Rule (RADAR) scores of two patients in the acute phase of first-episode depression, namely one with major dysmood disorder (MDMD) and another with simple dysmood disorder (SDMD). ACE: Adverse childhood experiences, AEs: adverse events; G-CoDe: general cognitive decline; SCIs: subjective cognitive impairments; SBs: suicidal behaviors; phenome1: first PC extracted from all symptom domains; phenome2: same as phenomen1 but includes SBs; phenome3: same as phenome2 but includes G-CoDe; LT: lifetime

**Figure 6.**
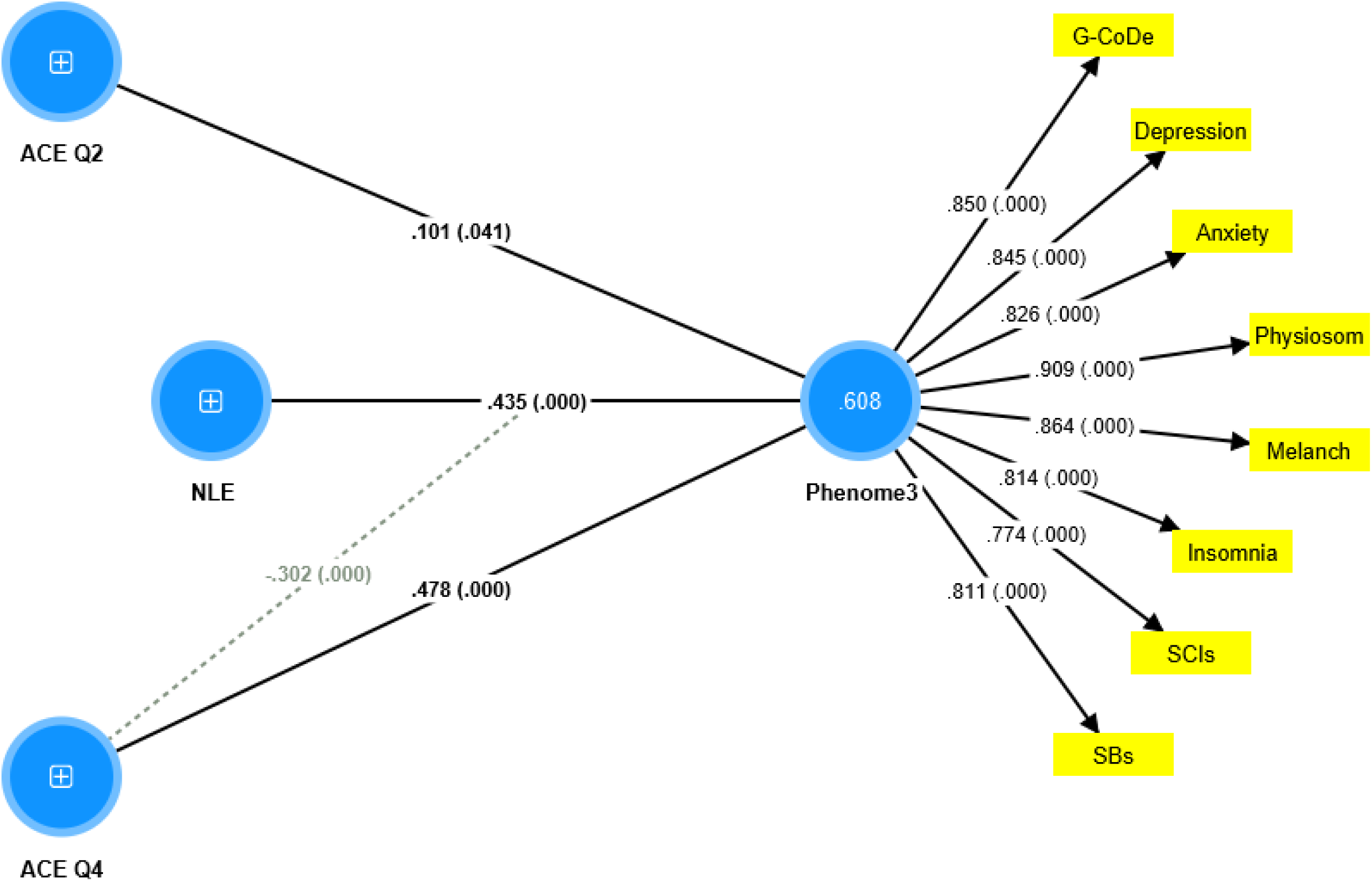
Results of Partial Least Squares (PLS) Analysis. ACE: adverse childhood experiences; NLE: negative life events; Q4: emotional neglect; Q2: physical abuse; G-CoDe: generalized cognitive decline; SCIs: subjective cognitive impairments; SBs: suicidal behaviors. Shown are pathway coefficients with exact p-values and loadings on the phenome factor.

### Results of PLS analyses

A first PLS model that considered the latent vector extracted from the phenome2 features as dependent variable, and the G-CoDe, AEs, and ACEs as explanatory variables. Moreover, the G-CoDe was entered as a mediating variable that was allowed to mediate the effects of ACE and AEs on the phenome2 score. With an SRMR of 0.051, the model quality fit was more than adequate. The convergent reliability was more than adequate for the phenome2 score (0.733) and the G-CoDe (0.609). The composite reliabilities of both constructs were more than adequate, namely 0.929 for the phenome2 score and 0.746 for the G-CoDe. PLSPredict shows that all Q^2^ predict values of the manifest and latent variables are positive indicating that the model outperforms the most naïve benchmark. Application of the CVPAT framework in PLSPredict examined the predictive reliability of the two endogenous constructs and shows that they have significantly lower average loss (t=3.04, p=0.003) and, thus, higher predictive validity than the indicator-average predictor benchmark. Nevertheless, the model is not really valid as no discriminatory validity is obtained: the HTMT ratio of 0.993 shows that the G-CoDe cannot be discriminated from the phenome2 score. Consequently, we have built a new model including the G-CoDe in the phenome3 factor as shown in **Figure 7**. The model fit was adequate with an SRMR of 0.042. The convergent and composite reliabilities of the pheome2 score were more than adequate with an AVE=0.701, rho_A = 0.944 and Cronbach alpha=0.939. PLSPredict shows that all manifest and latent variable Q2 values are positive. CVPAT shows that the average loss differences of PLS-SEM versus the indicator average (t=5.07, p<0.001) and linear model (t=2.50, p=0.014) are significant indicating a strong (t=3.04, p=0.003) predictive validity of the construct. PLS path analysis performed using 5,000 bootstraps shows that 60.8% of the variance in phenome2 is explained by the regression on ACEQ2, ACEQ4 and NLE and that also the mediating effect (interaction Q4 and NLE) is significant and shows an inverse effect. PLS Multigroup analysis and permutation multigroup analysis shows no differences in the model parameters between men and women, and smokers and non-smokers.

**Figure 7.**
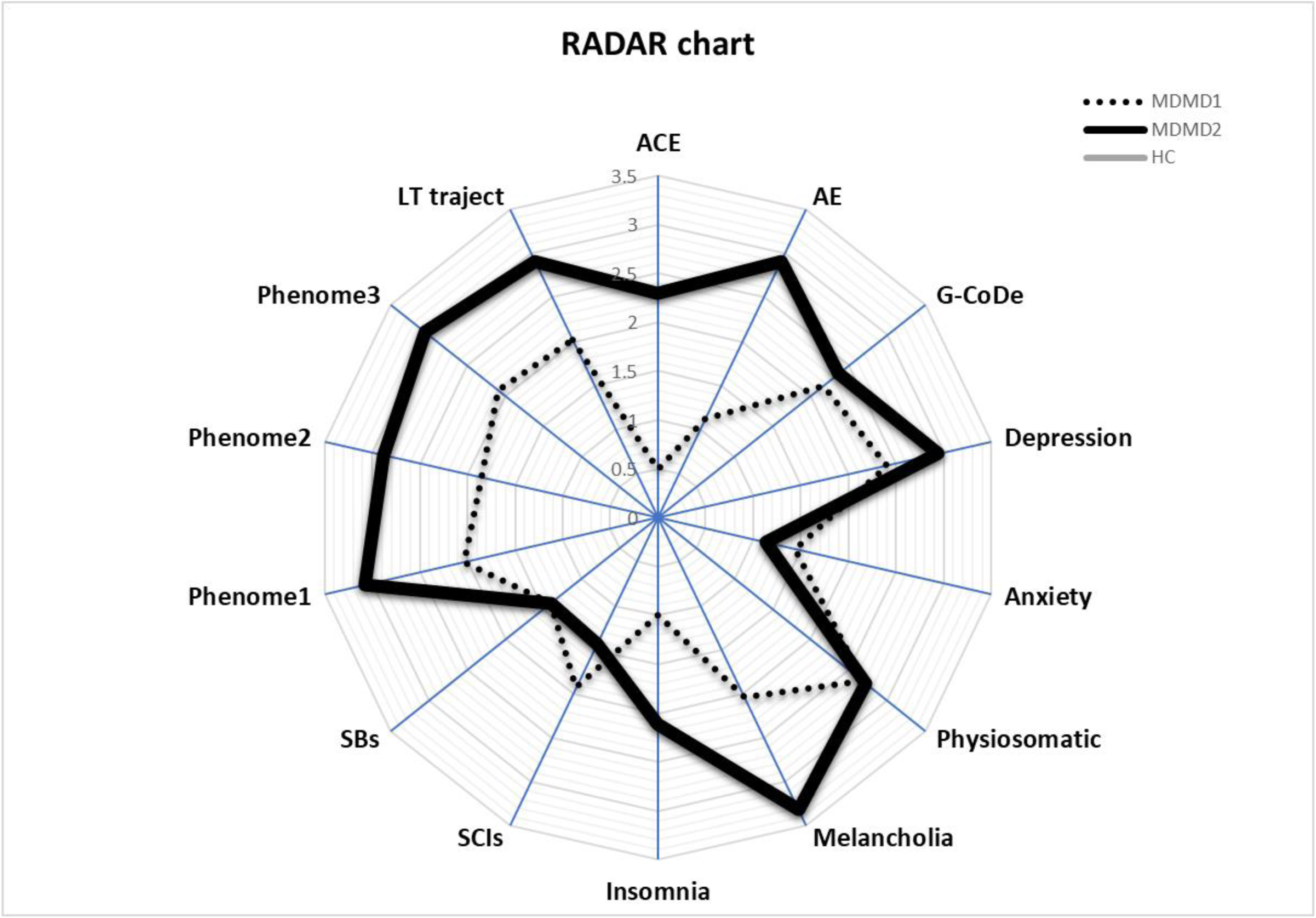
Radar or spider plot displaying the Research and Diagnostic Algorithmic Rule (RADAR) scores of two patients in the acute phase of first-episode major dysmood disorder (MDMD) ACE: Adverse childhood experiences, AEs: adverse events; G-CoDe: general cognitive decline; SCIs: subjective cognitive impairments; SBs: suicidal behaviors; phenome1: first PC extracted from all symptom domains; phenome2: same as phenomen1 but includes SBs; phenome3: same as phenome2 but includes G-CoDe; LT: lifetime

### Effects of the drug state

Since part of the first-episode patients were drug-naïve, we were able to decipher possible differences between drug-naïve and medicated patients. The latter were treated with antidepressant or atypical antipsychotics for at least 3 weeks. Introducing the medication status (drug-naïve versus use of psychotropic drugs) showed that there was no significant effect of the drug state on phenome3 (t=1.76, p=0.079). MICOM showed that the permutation p-values for the variables were non-significant indicating computational invariance when comparing drug-naïve and medicated patients. The mean original differences fell within the 2.5% and 97.5% limits suggesting invariance in composite equality. Consequently, we performed PLS MGA and permutation MGA. These analyses showed no significant differences in model parameters, including pathway coefficients and outer loadings (bootstrap MGA, parametric test, and Welch-Satterthwait test) and quality criteria (explained variance, AVE, etc) between drug-naive and medicated patients. **Figure 8** shows the differences in the RADAR scores between drug-naïve and medicated MDD patients. ANOVAs showed increased scores of pure depression (F=4.10, df=1/69, p=0.047), SBs (F=67.96, df=1/69, p<0.001), phenome2 (F=7.43, df=1/69, p=0.008) and phenome3 (F=6.59, df=1/69, p=0.012) in medicated as compared with drug-naïve patients. In contrast, sleep disorders were better in the medicated group (F=5.34, df=1/69, p=0.024).

**Figure 8.**
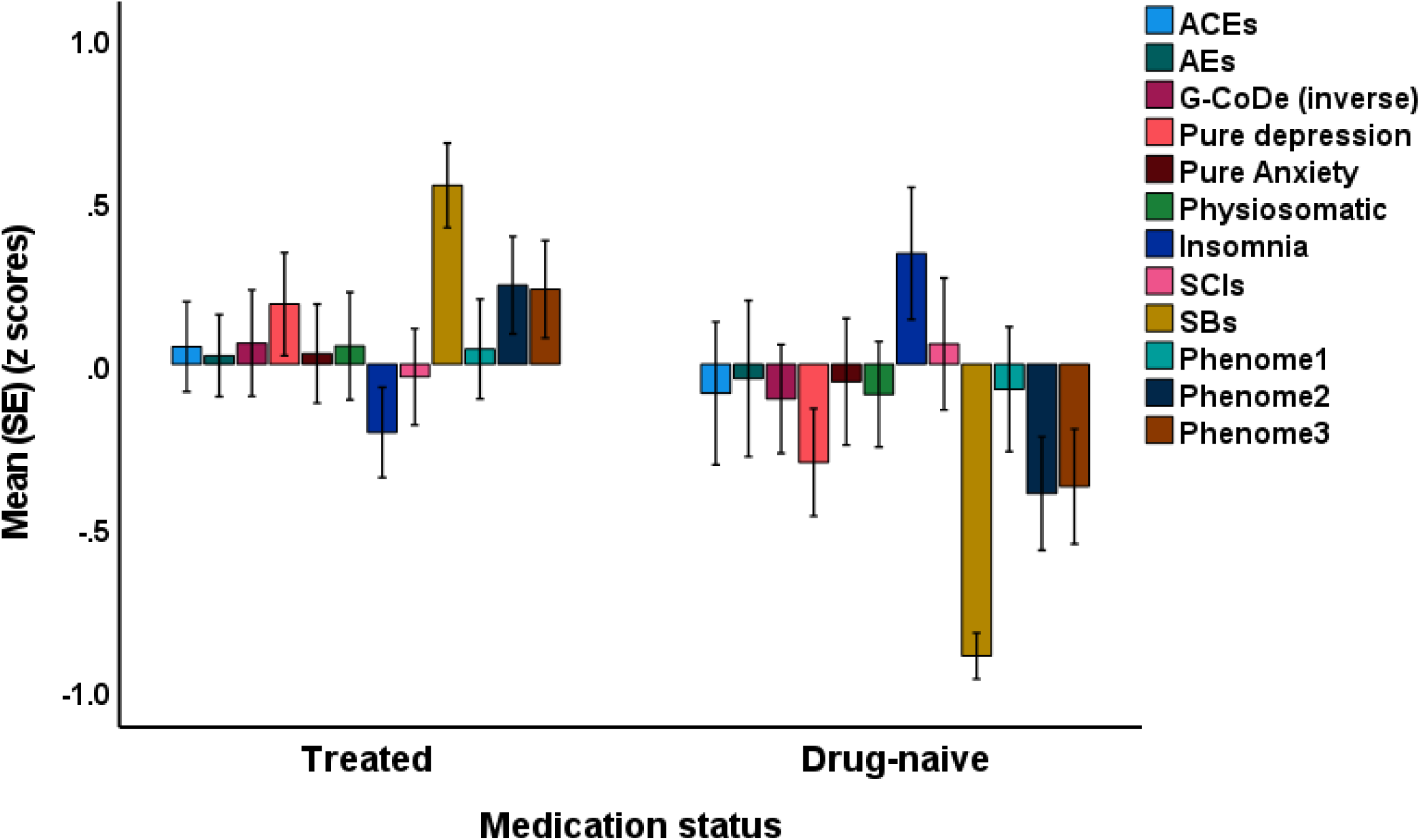
Clustered-bar graph showing the mean (SE) feature scores of the acute phase of first-episode depression divided into drug-naïve and treated patients. ACE: adverse childhood experiences; AEs: adverse events; G-CoDe: generalized cognitive decline; SCIs: subjective cognitive impairments; SBs: suicidal behaviors; LT traject: lifetime trajectory

### Features of first episode versus second episode MDD

Lastly, we have also examined differences between the 71 first-episode MDD patients and 19 second-episode MDD patients. **Table 4** shows the differences between both groups and healthy controls. We found that pure depressive symptoms and lifetime trajectory scores were significantly higher in second-episode than first-episode patients. Table 2 shows the associations between ACEs/AEs and the different RADAR scores in this study sample. Table 3 shows that a considerable part of the variance in the ROI score (55.5%) is explained by ACEQ4, NLE and the interaction pattern between ACEQ4 and NLE (negative effect).

**Table 4.**
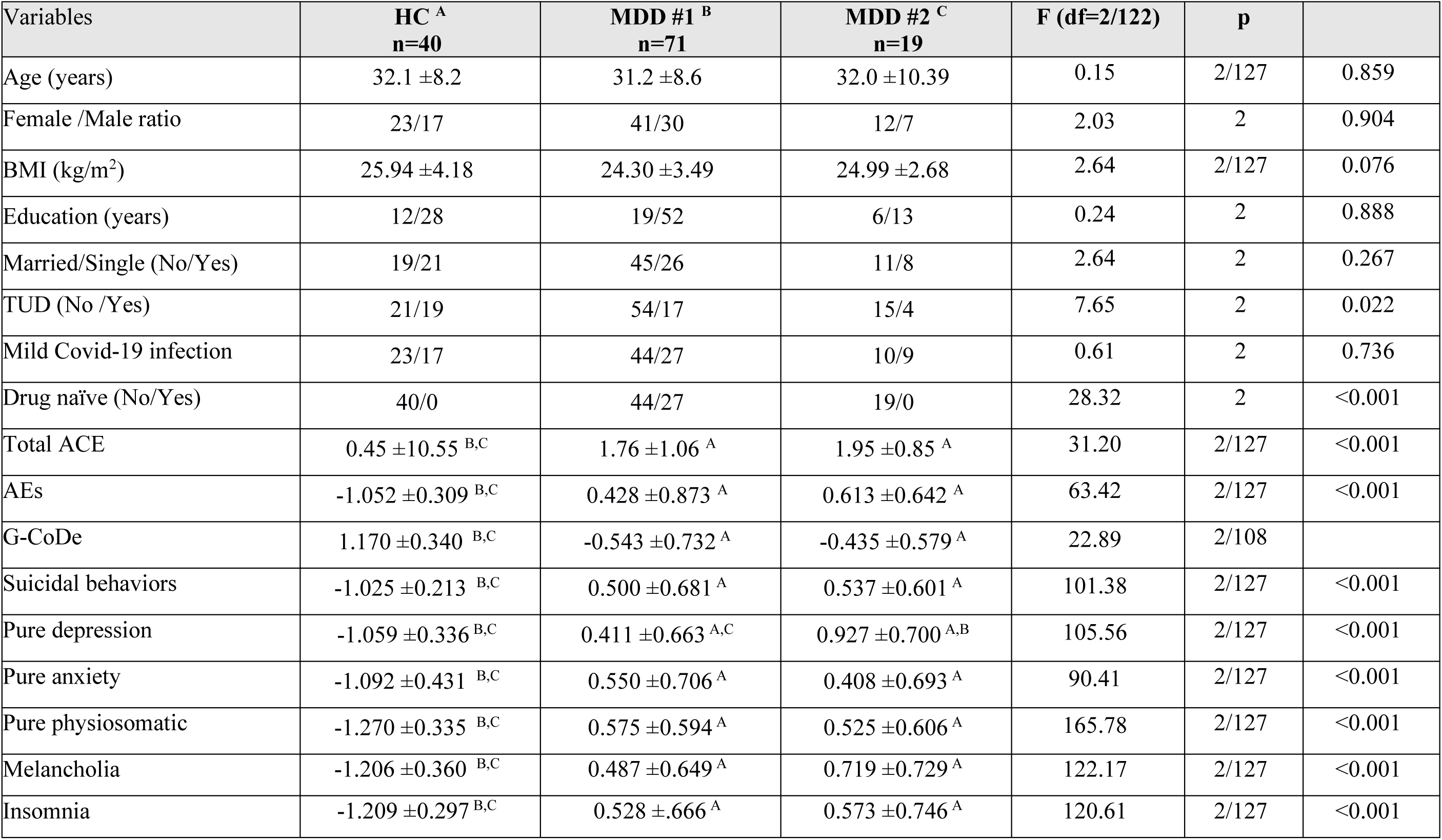
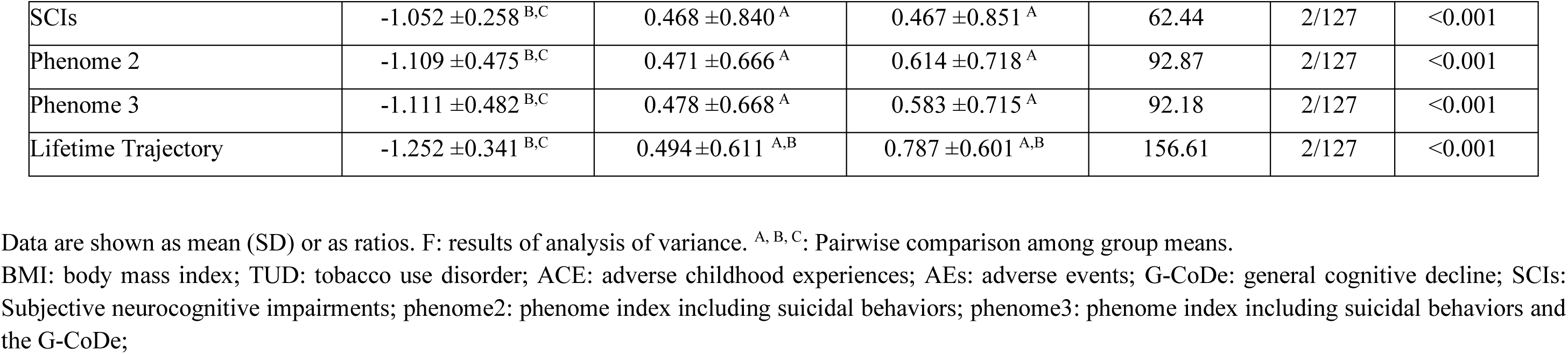
Features of healthy controls (HCs) and patients with major depressive disorder (MDD) who were divided into those with a first depressive episode (MDD #1) and a second depressive episode (MDD #2).

| Variables | HC <sup>A</sup><br>n=40 | MDD #1 <sup>B</sup><br>n=71 | MDD #2 <sup>C</sup><br>n=19 | F (df=2/122) | p |  |
| --- | --- | --- | --- | --- | --- | --- |
| Age (years) | 32.1 ±8.2 | 31.2 ±8.6 | 32.0 ±10.39 | 0.15 | 2/127 | 0.859 |
| Female /Male ratio | 23/17 | 41/30 | 12/7 | 2.03 | 2 | 0.904 |
| BMI (kg/m<sup>2</sup>) | 25.94 ±4.18 | 24.30 ±3.49 | 24.99 ±2.68 | 2.64 | 2/127 | 0.076 |
| Education (years) | 12/28 | 19/52 | 6/13 | 0.24 | 2 | 0.888 |
| Married/Single (No/Yes) | 19/21 | 45/26 | 11/8 | 2.64 | 2 | 0.267 |
| TUD (No /Yes) | 21/19 | 54/17 | 15/4 | 7.65 | 2 | 0.022 |
| Mild Covid-19 infection | 23/17 | 44/27 | 10/9 | 0.61 | 2 | 0.736 |
| Drug naïve (No/Yes) | 40/0 | 44/27 | 19/0 | 28.32 | 2 | <0.001 |
| Total ACE | 0.45 ±10.55 <sup>B,C</sup> | 1.76 ±1.06 <sup>A</sup> | 1.95 ±0.85 <sup>A</sup> | 31.20 | 2/127 | <0.001 |
| AEs | -1.052 ±0.309 <sup>B,C</sup> | 0.428 ±0.873 <sup>A</sup> | 0.613 ±0.642 <sup>A</sup> | 63.42 | 2/127 | <0.001 |
| G-CoDe | 1.170 ±0.340 <sup>B,C</sup> | -0.543 ±0.732 <sup>A</sup> | -0.435 ±0.579 <sup>A</sup> | 22.89 | 2/108 |  |
| Suicidal behaviors | -1.025 ±0.213 <sup>B,C</sup> | 0.500 ±0.681 <sup>A</sup> | 0.537 ±0.601 <sup>A</sup> | 101.38 | 2/127 | <0.001 |
| Pure depression | -1.059 ±0.336 <sup>B,C</sup> | 0.411 ±0.663 <sup>A,C</sup> | 0.927 ±0.700 <sup>A,B</sup> | 105.56 | 2/127 | <0.001 |
| Pure anxiety | -1.092 ±0.431 <sup>B,C</sup> | 0.550 ±0.706 <sup>A</sup> | 0.408 ±0.693 <sup>A</sup> | 90.41 | 2/127 | <0.001 |
| Pure physiosomatic | -1.270 ±0.335 <sup>B,C</sup> | 0.575 ±0.594 <sup>A</sup> | 0.525 ±0.606 <sup>A</sup> | 165.78 | 2/127 | <0.001 |
| Melancholia | -1.206 ±0.360 <sup>B,C</sup> | 0.487 ±0.649 <sup>A</sup> | 0.719 ±0.729 <sup>A</sup> | 122.17 | 2/127 | <0.001 |
| Insomnia | -1.209 ±0.297 <sup>B,C</sup> | 0.528 ±.666 <sup>A</sup> | 0.573 ±0.746 <sup>A</sup> | 120.61 | 2/127 | <0.001 |
| SCIs | -1.052 ± 0.258<sup>B,C</sup> | 0.468 ± 0.840<sup>A</sup> | 0.467 ± 0.851<sup>A</sup> | 62.44 | 2/127 | <0.001 |
| Phenome 2 | -1.109 ± 0.475<sup>B,C</sup> | 0.471 ± 0.666<sup>A</sup> | 0.614 ± 0.718<sup>A</sup> | 92.87 | 2/127 | <0.001 |
| Phenome 3 | -1.111 ± 0.482<sup>B,C</sup> | 0.478 ± 0.668<sup>A</sup> | 0.583 ± 0.715<sup>A</sup> | 92.18 | 2/127 | <0.001 |
| Lifetime Trajectory | -1.252 ± 0.341<sup>B,C</sup> | 0.494 ± 0.611<sup>A,B</sup> | 0.787 ± 0.601<sup>A,B</sup> | 156.61 | 2/127 | <0.001 |

Data are shown as mean (SD) or as ratios. F: results of analysis of variance. <sup>A, B, C</sup>: Pairwise comparison among group means.
BMI: body mass index; TUD: tobacco use disorder; ACE: adverse childhood experiences; AEs: adverse events; G-CoDe: general cognitive decline; SCIs: Subjective neurocognitive impairments; phenome2: phenome index including suicidal behaviors; phenome3: phenome index including suicidal behaviors and the G-CoDe;

## Discussion

### RADAR scores and RADAR plots

The first major finding of this study is that we were able to create RADAR scores and a RADAR plot in first-episode depression representing the phenome, SBs, G-CoDe, and lifetime trajectory scores. This verifies our prior studies indicating that such scores may be calculated for patients with recurrent mood disorders and that the scores can be displayed in a RADAR plot (Maes. 2022; 2023). Therefore, the latter enables the display of the mean differences in all features as standard deviations between MDMD, SDMD, and controls, as well as a patient-specific profile or fingerprint.

Firstly, the current study found that one factor could be extracted from different symptom domains (including pure depression, pure anxiety, physiosomatic, melancholic, insomnia, and SCIs), indicating that these distinct symptom profiles are manifestations of a shared or common core, i.e. the phenome of first-episode depression. This suggests that these symptom domains should be seen as highly intercorrelated domains driven by a shared pathophysiology underlying these phenome presentations. Sensitization in immunological and growth factor networks, activated oxidative stress pathways, decreased antioxidant defenses, enhanced bacterial translocation, and autoimmune responses are linked with phenome scores in recurrent unipolar and bipolar depression, as we have demonstrated previously (Maes et al., 2018; 2019; Simeonova et al., 2019; Maes, 2022; 2023).

Secondly, our results that SBs are part of the phenome of first-episode depression confirms our previous research on SBs that the latter can be expressed as RADAR scores which are a part of the phenome of recurrent depression and mood disorders (Maes, 2022; Maes, 2023). This again demonstrates that a same pathophysiology underlies both the phenome and SBs. Combinations of immune-inflammatory and nitro-oxidative pathways are related with suicidal ideation and suicidal attempts in people with mood disorders, according to two recent systematic reviews and meta-analyses (Vasupanrajit et al., 2021; 2022). In contrast to recurring MDD and mood disorders, our study on first-episode depression was unable to incorporate the ROI index, a key driver of SBs and the phenome of mood disorders (Maes et al., 2018; 2021; 2022a, Maes, 2022, 2023). In this regard, it is intriguing to notice that second-episode patients had higher pure depressive and lifetime trajectory ratings than first-episode patients. Thus, it is possible to conclude that the deteriorating influence of ROI on the phenome (Maes et al., 2019) is already present in the second episode.

Thirdly, we discovered that a general impairment in cognitive abilities during the acute phase of first-episode depression constituted an additional component of the phenome and, as such, should be considered a manifestation of the phenome of depression. In this work, we were able to extract one latent vector from the VFT, MMSE, and memory, judgment, and orientation tests, thereby demonstrating a G-CoDe in first-episode depression. Executive functioning, semantic and episodic memory, recall, strategy utilization, rule acquisition, emotional recognition, visual sustained attention, and attentional set-shifting were used to develop a G-CoDe for schizophrenia previously (Maes and Kanchanatawan, 2021). In contrast to these findings in stable phase schizophrenia, however, the current investigation indicated that the G-CoDe could not be distinguished from the phenome of acute depression, indicating that the G-CoDe is a feature of the acute phase’s severity. As a consequence, these impairments will probably normalize throughout the remitted and euthymic phases, given that inter-episode cognitive deterioration increases with ROI (Maes et al., 2019).

Overall, our method of using different RADAR scores to indicate the acute state of MDD contrasts with the current gold standard method of employing a binary diagnosis (“major depressive episode, single episode”). It is clear that different numeric RADAR values convey far more information of the actual state of depression than a binary diagnosis, which additionally is unreliable. In addition, based on the phenome, G-CoDe, and SB RADAR values, we were able to establish two groups, with one cluster exhibiting a very severe phenome, G-CoDe, SB, ACE, and AE scores, labeled first-episode “MDMD”, as opposed to a cluster of first-episode “SDMD” patients. Further research utilizing cross-validation approaches, such as statistical isolinear multiple component analysis (SIMCA), is required to determine whether the observed differences are qualitative or quantitative (Maes et al., 1990). The second question is whether this cluster of MDMD patients includes those who will eventually develop the phenotype characterized by high ROI scores, neuro-immune and neuro-oxidative stress biomarkers, and increased phenome scores (Maes et al, 2019; 2021; Maes, 2022; 2023).

It follows that our quantitative RADAR scores and the MDMD/SDMD diagnosis should be employed as dependent factors in regression or neural network studies with the neuro-immune and neuro-oxidative biomarkers as explanatory variables. Using a post-hoc, erroneous, and unreliable higher-order binary construct (the DSM/ICD diagnosis) as an explanatory variable in t-tests or ANOVAs with the biomarkers as dependent variables is grossly inaccurate and should be replaced by our method (Maes, 2022; 2023; Stoyanov and Maes, 2022). In actuality, the optimal strategy would be to calculate two types of RADAR scores, namely those of the acute (this study; Maes et al., 2022a) and residual (Maes et al., 2021) phases. Consequently, this technique will permit us to determine the state and trait biomarkers of SBs, ROI, and MDMD in comparison to SDMD and controls.

### ACEs, NLE and first episode depression

The second major finding of this study is that the number of ACEs and NLEs significantly predict G-CoDe, SBs and phenome scores. In earlier research, we discovered that ACEs have a significant effect on G-CoDe, ROI, SBs, and the acute and residual phase phenome of depression (Maes et al., 2018; 2022b). There is now compelling evidence that the cumulative effects of adverse childhood experiences (ACEs) are causally connected with the onset and severity of depression later in adulthood, as well as cognitive impairments and suicidal tendencies (Jansen et al., 2016; Hadland et al., 2012; Alvarez et al., 2011; Aas et al., 2017; Moraes et al., 2017; Schoedl et al., 2010; Miao et al., 2022; Whitaker et al., 2021; Pechtel and Pizzagalli, 2011). In a recent study involving patients from Brazil, we discovered that emotional abuse and neglect, physical neglect, and physical and sexual abuse (combined into one latent vector) were significantly linked with AOPs, ROI, cognitive deficits, and the phenome of depression (Maes et al., 2018; 2019). In Thai patients, we discovered that a latent vector derived from mental neglect and trauma, physical trauma, and domestic violence predicted the phenome and ROI of depression (Maes et al., 2022b). In the latter study, sexual abuse, and a factor from parental loss due to separation, death, or divorce, and a family history of mental illness were independently related with depression. This contradicts the current study’s conclusions that no latent vector could be identified from the 10 ACE scores included for Iraqi patients and that emotional neglect and physical abuse were the most important predictors.

In this work, we determined that NLEs have a significant influence on G-CoDe, SBs, and the phenome of first-episode depression, above and beyond the effects of ACEs. It is known that NLEs are connected with the onset of depression, including depressive symptoms in students and late-life depression, according to a large body of research (Paykel, 2003; Kendler et al., 1999; Kraaij et al., 2022; Ji L et al., 2021; Guang et al., 2017). Furthermore, there is evidence that stressful life events may produce depression, rather than the other way around (Phillips et al., 2015). In addition, major life events play a greater role in the development of first-onset depression than in subsequent episodes (Paykel., 2003).

Moreover, in the present investigation, we discovered that ACEs and, in particular, emotional neglect may negatively moderate the influence of NLEs. This suggests that although ACEs and NLEs have cumulative effects on the phenome, there is also a substantial interaction effect indicating that increased emotional neglect may reduce the influence of NLEs, suggesting that the impact of the latter may be capped as ACEs increase. There are currently indications that the effects of ACEs on the phenome, including SBs, are mediated by sensitization of the cytokine and growth factor networks, which are reactivated in response to new immunological stimuli (Maes et al., 2022b). There is also evidence that psychological stressors can activate T helper-1 and M1 macrophage cytokine networks in humans (Maes et al., 1998; 1999a; 1999b; Gu et al., 2012; Priyadarshini and Aich, 2012). Based on the present study’s findings that ACEs and NLEs have cumulative effects on the phenome, we may hypothesize that NLEs may operate as a second hit to reactivate the sensitized cytokine network. However, our findings imply that this effect is dependent on the amount of ACEs, such that a greater number of ACEs may be associated with a diminished influence of NLEs. Since there is a strong association between ACEs and NLEs, it may also be that people with ACEs select environments with more stressful events and distress and violence, explaining that part of the links between NLEs and the phenome may be non-causal (Kendler et al., 1999).

### Limitations

If we had assessed the sensitization of the immune system utilizing LPS+PHA-stimulated production of various cytokines and growth factors, this study would have been incredibly intriguing (Maes et al., 2022a). As we have now produced RADAR scores and plots for the acute period of first-episode depression, recurrent depression, and the remitted phase of depression in Iraqi, Thai, and Brazilian patients, future research should establish these scores in Western and Caucasian study groups. Another potential complicating aspect is the medication state of some patients. Nevertheless, after including the impact of ACEs and NLEs in the PLS analysis, the drug state of the patients had no effect on the phenome, as determined by regression analysis. Importantly, when comparing drug-naive individuals with treated first-episode patients, the latter demonstrated greater (albeit with small effect sizes) SB, pure depression, and phenome ratings, whereas the former demonstrated superior sleeplessness. Probably these results show that people with suicidal behaviors are more pro-actively treated with psychotropic medications. Another possible limitation is the putative inference with the effects of acute COVID infection which may cause Long COVID with depression, anxiety, chronic fatigue and physiosomatic symptoms (Al-Hakeim et al., 2022). Nevertheless, this Long COVID phenome is predicted by critical disease during the acute phase with high fever and low oxygen saturation, whilst this study excluded COVID-19 patients who had suffered from moderate or critical COVID disease.

## Conclusions

In this study, we demonstrated how to develop a validated precision model, RADAR scores, and RADAR plots for first-episode depression patients. We found that ACEs and NLEs are related to SBs, cognitive impairments, and first-episode depression symptoms. We discovered that one phenome factor underpins depressive, anxious, fatigue, physiosomatic, and melancholic symptoms, insomnia, SB, and cognitive impairments. Importantly, 60.8% of the variance in this phenome’s RADAR score was explained by the cumulative effects of ACEs and NLEs, as well as the pattern of interaction between emotional neglect and NLEs, which suggests that ACEs may attenuate the effects of NLEs on the phenome. Future research should associate biomarkers with the new RADAR scores and MDMD diagnosis computed during the acute and partially remitted phases, instead of using the binary diagnosis of MDD. Moreover, it would be more interesting to use RADAR graphs in clinical practice than an unreliable DSM or ICD diagnosis because the former provides more accurate information and additionally provides a personalized fingerprint of the patient’s status.

## Declarations

### Ethics approval and consent to participate

All participants provided written informed consent prior to the enrollment in the study. The study complied with international and Iraqi laws governing ethics and privacy. According to the International Guidelines for the Protection of Human Subjects of the Declaration of Helsinki, the study was approved by the ethics committee (IRB) of the College of Medical Technology, The Islamic University of Najaf, Iraq (Document No. 18/2021).

### Availability of data and materials

The dataset (PLS models) generated during and/or analyzed during the current study will be available from Prof. Dr. Michael Maes upon reasonable request and once the dataset has been fully exploited by the authors.

## Acknowledgements

We gratefully acknowledge the help of all psychiatry staff involved in the execution of this study.

